# Acute dorsal genital nerve stimulation increases subjective arousal in women with and without spinal cord injury: a preliminary investigation

**DOI:** 10.1101/2023.04.24.23288935

**Authors:** Elizabeth C. Bottorff, Priyanka Gupta, Giulia M. Ippolito, Mackenzie B. Moore, Gianna M. Rodriguez, Tim M. Bruns

## Abstract

**Introduction:** Female sexual dysfunction (FSD) impacts an estimated 40% of women. Unfortunately, female sexual function is understudied, leading to limited treatment options for FSD. Neuromodulation has demonstrated some success in improving FSD symptoms. We developed a pilot study to investigate the short-term effect of electrical stimulation of the dorsal genital nerve and tibial nerve on sexual arousal in healthy women, women with FSD, and women with spinal cord injury (SCI) and FSD.

**Methods:** This study consists of a randomized crossover design in three groups: women with SCI, women with non-neurogenic FSD, and women without FSD or SCI. The primary outcome measure was change in vaginal pulse amplitude (VPA) from baseline. Secondary outcome measures were changes in subjective arousal, heart rate, and mean arterial pressure from baseline. Participants attended one or two study sessions where they received either transcutaneous dorsal genital nerve stimulation (DGNS) or tibial nerve stimulation (TNS). At each session, a vaginal photoplethysmography sensor was used to measure VPA. Participants also rated their level of subjective arousal and were asked to report any pelvic sensations.

**Results:** We found that subjective arousal increased significantly from before to after stimulation in DGNS study sessions across all women. TNS had no effect on subjective arousal. There were significant differences in VPA between baseline and stimulation, baseline and recovery, and stimulation and recovery periods among participants, but there were no trends across groups or stimulation type. Two participants with complete SCIs experienced genital sensations.

**Discussion:** This is the first study to measure sexual arousal in response to acute neuromodulation in women. This study demonstrates that acute DGNS, but not TNS, can increase subjective arousal, but the effect of stimulation on genital arousal is inconclusive. This study provides further support for DGNS as a treatment for female sexual dysfunction.

## Introduction

Female sexual function has been historically understudied, leading to limited treatment options for the approximately 40-50% of women who suffer from symptoms associated with female sexual dysfunction (FSD)^1^. Existing treatment options for FSD, such as bremelanotide^2^ and flibanserin^3^, primarily target hypoactive sexual desire disorder (HSDD). There is a lack of treatments that target challenges with the physiological aspects of sexual function, such as lubrication or arousal. Sildenafil, a successful pharmaceutical for treating male sexual arousal dysfunction, was pursued for FSD but ultimately abandoned due to its low efficacy rate and high incidence of adverse events^4^.

FSD can have a variety of etiologies, one of which is spinal cord injury (SCI). People with SCI report sexual function as one of their top priorities to regain^5,6^ and sexual function is an important factor in quality of life for all adults^7^. Location and severity of the injury often determine which aspects of sexual function are impacted (e.g., arousal, desire). Psychogenic, but not reflexogenic, arousal is often possible in women with sacral level injuries while reflexogenic arousal is generally retained in women with injuries above the lumbar level^8^. These two examples demonstrate the heterogeneity in FSD symptoms among women with SCI, an underserved population that would particularly benefit from FSD treatments developed with pathophysiology taken into consideration.

Neuromodulation, or electrical stimulation of neural targets, has shown some promise in treating FSD in non-neurogenic women. Clinical trials using sacral neuromodulation to treat women with bladder dysfunction found that their sexual function, as evaluated by the female sexual function index (FSFI)^9^, improved as an unanticipated benefit^10–12^. Other bladder dysfunction neuromodulation targets have been investigated as treatments for FSD, including tibial nerve stimulation (TNS)^13,14^ and dorsal genital nerve stimulation (DGNS)^14^. Although the mechanisms of these interventions are not fully understood, we theorize that DGNS and TNS can improve FSD by increasing genital arousal. Genital arousal, often measured by vaginal blood flow, has shown to have short-term increases during peripheral nerve stimulation in preclinical models. Animal studies using TNS have shown increases in vaginal blood flow^15,16^ and we hypothesize that the underlying mechanisms involve a spinal reflex pathway. Similarly, animal studies using pudendal nerve stimulation, the proximal source of the dorsal genital nerve, have shown increases in vaginal blood flow^17,18^. These studies suggest that pudendal nerve stimulation activates spinal pathways that in turn activate the pelvic efferents that modulate vaginal blood flow. We hypothesize that increased blood flow contributes, at least in part, to improved FSFI scores for women with FSD.

Clinical studies on female sexual function often measure genital arousal, however this is the first study to measure genital arousal in response to transcutaneous neuromodulation. We sought to investigate if acute neuromodulation can modulate vaginal blood flow in women with SCI, able-bodied women with non-neurogenic FSD, and able-bodied women without FSD as healthy controls. Our goal was to assess if acute transcutaneous neuromodulation of the dorsal genital or tibial nerve can modulate genital and subjective arousal. We chose these three groups of participants to assess which treatments were best at evoking a blood flow response and subjective arousal given the presence of SCI or FSD.

## Materials and Methods

All study activities were approved by the University of Michigan Institutional Review Board (HUM00148746) prior to initiation and all data was collected at Michigan Medicine between November 2020 and March 2022. We recruited participants via physician referral, flyers placed in relevant clinics in the local area, and online through a University of Michigan health research portal. The study is registered at clinicaltrials.gov under identifier NCT04384172.

This study consists of a randomized crossover design with three groups: women with SCI (SCI), women with non-neurogenic FSD (FSD), and women with No Dysfunction and who are Able-Bodied (NDAB). Participants were screened for eligibility with a clinical study coordinator prior to enrollment. All participants were over 18 years old, biologically female, and sexually active at least once a month. SCI participants could be interested in sexual activity if not sexually active. To be included in the SCI arm, participants had to have a clinically diagnosed spinal cord injury at grade AIS (American Spinal Injury Association Impairment Scale) A-C at a level within C6-S1 at least six months prior to enrollment and a short-form FSFI^19^ score below 19. Women with FSD were neurologically intact with a short-form FSFI score below 19 and an FSFI lubrication sub-score below or equal to 3. Women without FSD were neurologically intact with a short-form FSFI score above or equal to 19 and FSFI lubrication sub-score above 4. Exclusion criteria for all participants were as follows: (1) pregnant, (2) clinically diagnosed bladder dysfunction, pelvic pain, or other pelvic organ symptoms, (3) active infection or active pressure sores in the perineal region, (4) epilepsy, and (5) implanted pacemaker or defibrillator. Additional exclusion criteria for SCI participants included worsening in motor or sensory function in the last month. NDAB and FSD participants were also excluded if they had clinically diagnosed bladder dysfunction, pelvic pain, or other pelvic organ symptom.

After obtaining informed consent, we instructed participants to submit demographic information and complete five clinically validated surveys: the American Urological Association Symptom Index (AUASI) bladder symptom index^20^, the female sexual function index (FSFI)^9^, the fecal incontinence severity index (FISI)^21^, the patient assessment of constipation-symptoms (PAC-SYM)^22^, and the short-form qualify of life survey (SF-36)^23^. The surveys were collected online in REDCap, a standard clinical tool for survey data collection^24^. Participants completed one or two study sessions corresponding to two stimulation targets: the dorsal genital nerve and tibial nerve. We used block randomization, with block sizes of 10 for each group, to determine which nerve target was used in the first study session. Study team members and participants were not blinded. Participant’s second study session was one to five months after their first. Participants filled out pelvic function surveys that asked them about their bladder, bowel, and sexual function for a given day. The surveys were filled out daily from two days prior to two days after each study session to monitor any carryover effects from neuromodulation.

At each study session, participants were asked to sit, partially reclined in a comfortable position. A vaginal photoplethysmography transducer (TSD204 Biopac Systems Inc., Goleta, CA) was placed in the vaginal canal to monitor vaginal pulse amplitude (VPA), a measurement of relative vaginal blood flow^25^. A clinician placed two round surface electrodes (1.25 inch diameter, ValuTrode Neurostimulation Electrodes CF3200, Axelgaard Manufacturing Co. Ltd., Fallbrook, CA) on either side of the clitoris^26^ for DGNS study sessions and above the malleolus and on the bottom of the foot^27^ for TNS study sessions. Stimulation was delivered with a transcutaneous electrical nerve stimulation (TENS) device (Empi Select 199584, Medi-Stim Inc., Wabasha, MN, USA). The stimulation amplitude was determined by slowly increasing it from 0 mV until a maximum comfortable level or 60 mV peak-to-peak was reached, whichever was lower. We recorded VPA at a sampling rate of 200 Hz during a 5-minute baseline period, 20 minutes of nerve stimulation at 20 Hz, 400 μs pulse width at the pre-determined amplitude, and a 5-minute post-stimulation recovery period for a total of 30 minutes. We asked participants to rate their level of subjective arousal on a 5-point Likert-style scale at four times throughout the recording trial: before baseline, before stimulation, after stimulation, and after the recovery period. After the trial, we asked participants their opinion of the TENS device, if it elicited any genital sensations, and if they would consider using it.

All data analysis was performed in MATLAB (Mathworks, Natick, MA, USA). VPA signals for each participant session were processed before subsequent analysis and statistics across participants. We bandpass filtered the raw VPA signal from 0.5 to 30 Hz and identified peaks and troughs using MATLAB’s findpeaks function. We visually inspected the peaks and troughs for artifact removal. We removed obvious artifacts if they did not conform to the typical sawtooth shape^28^ or had a trough-to-peak amplitude that had over a 100% increase from the previous waveform^29^. The median percentage of data points removed was 5.5% and ranged from 1.3% to 51.5%. We calculated trough to peak amplitude and binned the data into 10 second intervals^30^. Binned values were averaged for three time periods: 5-minute baseline (VPA_Baseline_), 20-minutes of stimulation (VPA_Stim_), and 5-minute recovery (VPA_Recovery_). We made comparisons between these three periods within each participant with a one-way Analysis of variance (ANOVA) followed by post-hoc pairwise Tukey HSD tests. We calculated the percent change for each participant between each of VPA_Baseline_, VPA_Stim_, and VPA_Recovery_, and made comparisons across participants for VPA_Change_ (VPA_Stim_ – VPA_Baseline_) for each stimulation location with a Wilcoxon signed rank test. We compared subjective arousal scores between each timepoint with a Wilcoxon signed rank test. We compared baseline heart rate and mean arterial blood pressure to the last heart rate and blood pressure recorded during stimulation with Wilcoxon signed rank tests. We compared the survey scores (SF-36, AUASI, PAC-SYM, FISI, and FSFI) across participants from the three different groups with Wilcoxon rank sum tests. All statistical analysis used alpha = 0.05 to determine significance.

## Results

We screened 101 participants for eligibility over the phone, of which 92 were either excluded, declined to participate, or were lost to follow-up. Ultimately, 3 participants in each group completed at least one study session. Participants in each group were lost to follow-up or became ineligible after the first session, leading to a total of five participants who completed two sessions. Figure 1 provides a breakdown of participant recruitment and retention. Demographics for all three groups of participants can be found in Table 1. Survey results averaged across each participant group can be found in Table 2. The survey scores were not significantly different between participant groups. The mean and standard error for stimulation amplitude for DGNS and TNS sessions was 28.8 ± 9.5 mV and 33.4 ± 10.0 mV peak to peak, respectively. Stimulation amplitudes for all sessions can be found in Tables 4 and 5.

**Figure 1.**
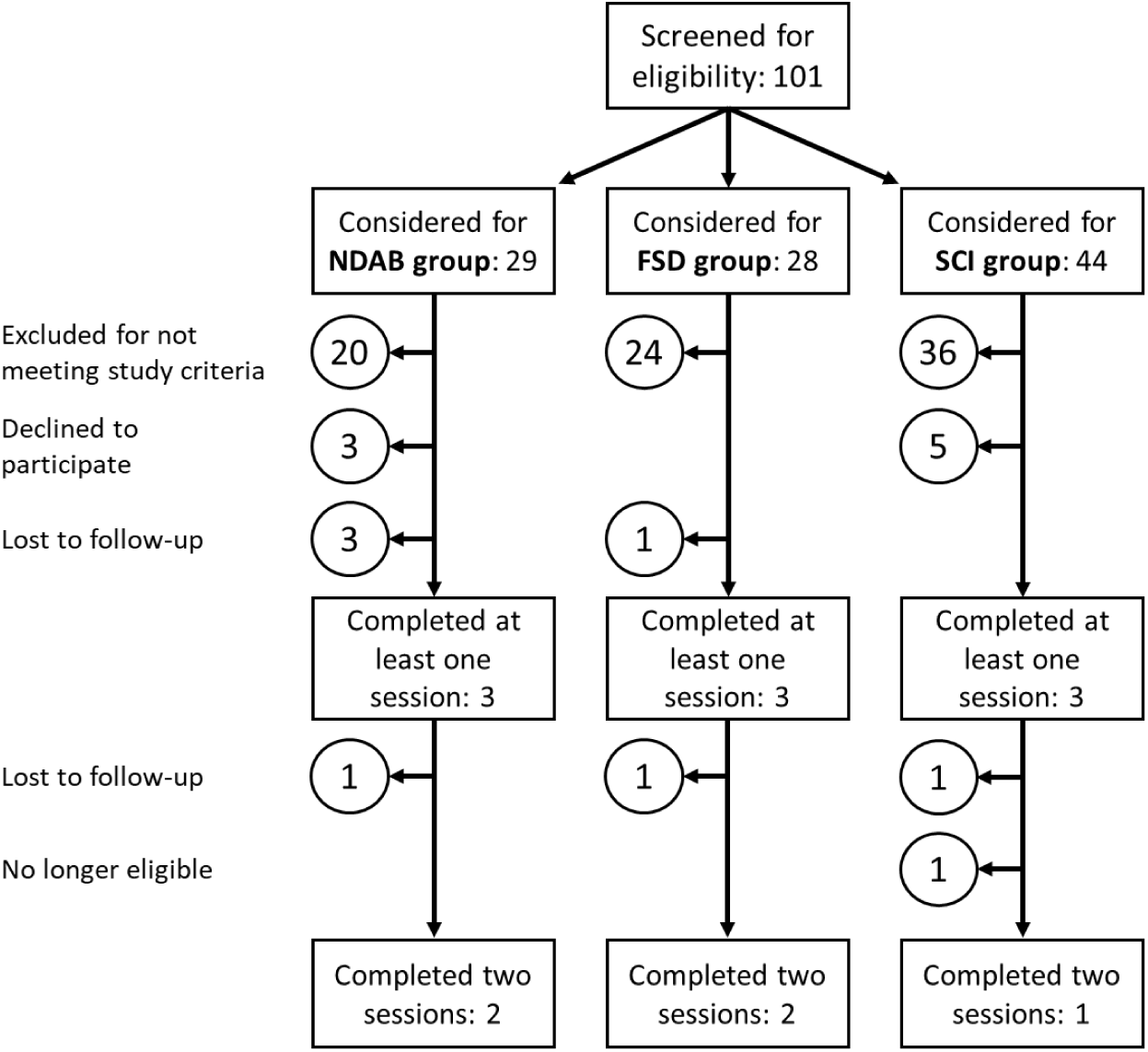
Participant screening and recruitment.

**Table 1.** Participant Demographics.

| Participant ID | Age | Height (m) | Weight (kg) | Race | Ethnicity |
| --- | --- | --- | --- | --- | --- |
| NDAB-1 | 21-25 | 1.60 | 64 | Asia or Pacific Islander | Non- Hispanic or Latino |
| NDAB-2 | 21-25 | 1.57 | 81 | White, Caucasian | Non- Hispanic or Latino |
| NDAB-3 | 26-30 | 1.52 | 77 | Multiracial | Hispanic or Latino |
| SCI-1 (sacral-level spina bifida presents like SCI) | 46-50 | 1.42 | 57 | White, Caucasian | Non- Hispanic or Latino |
| SCI-2 (T5, AIS-A, 23 months post-injury) | 46-50 | 1.60 | 68 | White, Caucasian | Non- Hispanic or Latino |
| SCI-3 (T2, AIS-A, 15 years post-injury) | 36-40 | 1.60 | 66 | White, Caucasian | Non- Hispanic or Latino |
| FSD-1 | 31-35 | 1.70 | 59 | White, Caucasian | Non- Hispanic or Latino |
| FSD-2 | 21-25 | 1.68 | 75 | Multiracial | Non- Hispanic or Latino |
| FSD-3 | 31-35 | 1.65 | 48 | White, Caucasian | Non- Hispanic or Latino |

**Table 2.** Participant Survey Results.

| Participant ID | SF36<br>(0 to 100+) | AUASI<br>(35 to 0+) | PACSYM<br>(48 to 0+) | FISI<br>(61 to 0+) | FSFI%<br>(2 to 36+) | FSFI<br>Lubrication<br>(1 to 6+) |
| --- | --- | --- | --- | --- | --- | --- |
| NDAB-1 ● | 76.9 | 2 | 2 | 0 | 32.5 | 6 |
| NDAB-2 ○ | 69.2 | 1 | 11 | 0 | 33.9 | 6 |
| NDAB-3 ■ | 53 | 5 | 0 | 3 | 23.5<sup>\$</sup> | 6 |
| SCI-1 □ | 35.5 | 8 | 10 | 13 | 22.7 | 3.9 |
| SCI-2 ▲ | 53.4 | 2 | 6 | 19 | 15.4 | 1.2 |
| SCI-3 Δ | 52.6 | 2 | 3 | 0 | 21.8 | 1.8 |
| FSD-1 ◆ | 74.6 | 0 | 0 | 0 | 12 | 1.2 |
| FSD-2 ◇ | 53.8 | 5 | 14 | 21 | 7.7 | 1.2 |
| FSD-3 ★ | 57.5 | 0 | 0 | 0 | 9.8 | 3 |
| Average NDAB | 66.4 | 2.7 | 4.3 | 1.0 | 30.0 | 6.0 |
| Average SCI | 47.2 | 4.0 | 6.3 | 10.7 | 20.0 | 2.3 |
| Average FSD | 62.0 | 1.7 | 4.7 | 7.0 | 9.8 | 1.8 |

+ indicates “no dysfunction” score
% clinical cutoff score for dysfunction = 26.55<sup>46</sup>
\$ low score due to temporary sexual inactivity after initial screening with short-form FSFI

In DGNS trials, we found significant (p = 0.0078) increases in subjective arousal from before the trial to after the stimulation period across all participants (Figure 2a). These changes were also significant (p = 0.0078) from before the trial until after the recovery period. Participants in each of the three groups reported increased arousal during DGNS. There were no significant changes in subjective arousal across TNS trials (Figure 2b). There were no significant differences between heart rate or mean arterial blood pressure between baseline and stimulation in DGNS or TNS trials. The daily pelvic function surveys indicated that most participant’s bladder, bowel, and sexual function were stable and no participants reported carry-over effects from the stimulation session.

**Figure 2.**
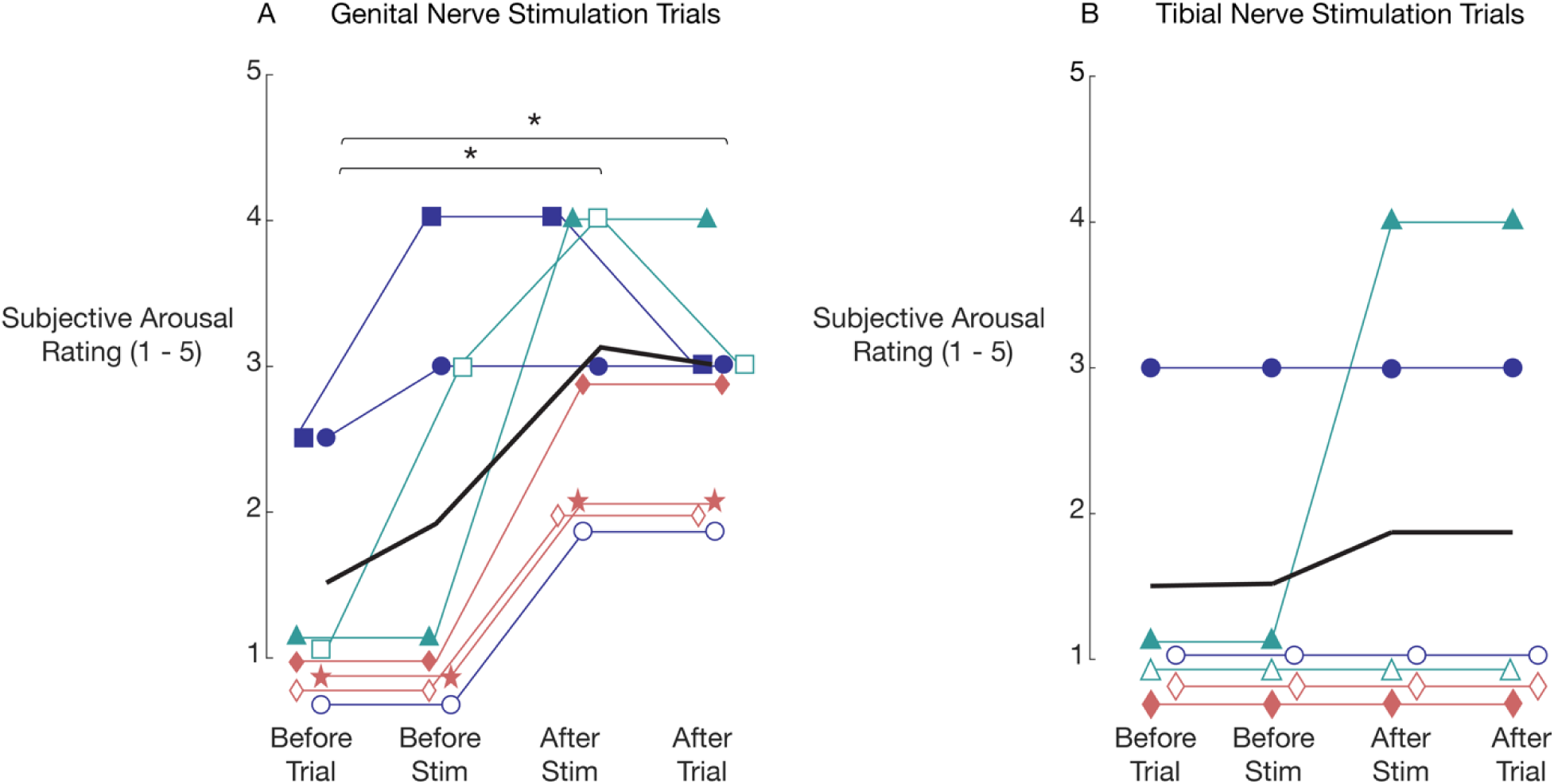
Subjective arousal scores for dorsal genital nerve stimulation (DGNS) and tibial nerve stimulation (TNS) sessions. Icons represent the same individual participants, as defined in Table 2. Icon color corresponds to group (indigo = able bodied, non-dysfunction, teal = SCI, rose = FSD). Solid black line denotes average values. Subjective arousal increased significantly (p = 0.0078) in DGNS trials, but not TNS trials.

Most participants reported genital sensations in response to DGNS (7/8). Comments about receiving DGNS were mostly neutral and participants indicated that they would be willing to use a TENS device at the dorsal genital nerve. Half of participants (3/6) reported genital sensations during TNS and seemed just as willing to use TENS at the tibial nerve if they knew it would help with their sexual dysfunction. Table 3 contains all participant feedback.

**Table 3.** Paraphrased participant responses to session follow up questions.

| Participant ID | Participant comments about DGNS |  |  | Participant comments about TNS |  |  |
| --- | --- | --- | --- | --- | --- | --- |
|  | What is your general opinion of the device? | Did it elicit any genital arousal responses (e.g., lubrication, tingling, pleasurable sensations)? | Would you consider further use of the device? | What is your general opinion of the device? | Did it elicit any genital arousal responses (e.g., lubrication, tingling, pleasurable sensations)? | Would you consider further use of the device? |
| NDAB-1 | Not painful | Lubrication and tingling | Yes | No discomfort, fine | Maybe lubrication | If dysfunction |
| NDAB-2 | Weird, kind of strange | Maybe lubrication towards the end of the stimulation | NA* | Numbing | No | No |
| NDAB-3 | Neutral, kind of strange | A little tingling | If had dysfunction | Lost to follow up |  |  |
| SCI-1 | Fine, comfortable | Lubrication, tingling, a little bit of pleasurable sensations | Yes | Lost to follow up |  |  |
| SCI-2 | It works, nothing uncomfortable about it. Has to concentrate to feel things | Tingling and pulsation | Yes | Easy at home, comfortable | Tingling and throbbing | Yes |
| SCI-3 | Withdrawn after becoming ineligible |  |  | Would use the device | Tingling and bladder spasms | Yes |
| FSD-1 | Neutral | Tingling | Yes | Neutral, ambivalent | No | Yes |
| FSD-2 | Pretty neutral | None, just felt like tapping | Might in a non-clinical setting | Fine | No | Potentially |
| FSD-3 | More warmth than arousal | Warmth and blood flow, but no arousal | Probably not | Lost to follow up |  |  |

\* NA denotes "Not Applicable" as participant misunderstood the question

VPA recordings from one participant in the NDAB group had corrupted data from both of their study sessions. All participants except one (FSD-3) had significant differences in their peak-to-peak VPA between at least two time periods in both DGNS and TNS sessions. These changes were not consistent within or across groups. Across DGNS sessions, two participants had an overall increase from VPA_Baseline_ to VPA_Recovery_ (NDAB-3, p < 0.001; SCI-1 p < 0.001), four had an overall decrease (NDAB-2, p < 0.001; SCI-2, p < 0.001; FSD-1, p = 0.009; FSD-2, p < 0.001), and one participant had no changes (FSD-3) (Figure 3). Across TNS sessions, 3 participants had an overall increase from VPA_Baseline_ to VPA_Recovery_ (NDAB-2, p < 0.001; FSD-1, p = 0.018; FSD-2, p < 0.001), 1 participant had an overall decrease (SCI-2, p = 0.004), and 1 participant had a decrease from VPA_Baseline_ to VPA_Stim_ that returned to baseline values in VPA_Recovery_ (SCI-3) (Figure 4). The average VPA_Change_ was +3.5 ± 26.7% and +3.5 ± 14.0% for DGNS and TNS sessions respectively, which were not significantly different from zero. Average VPA_Baseline_, VPA_Stim_, and VPA_Recovery_, as well as percent changes between each of the time periods for each participant, can be found in Tables 4 & 5. Subjective arousal and VPA data are publicly accessible online^31^.

**Figure 3.**
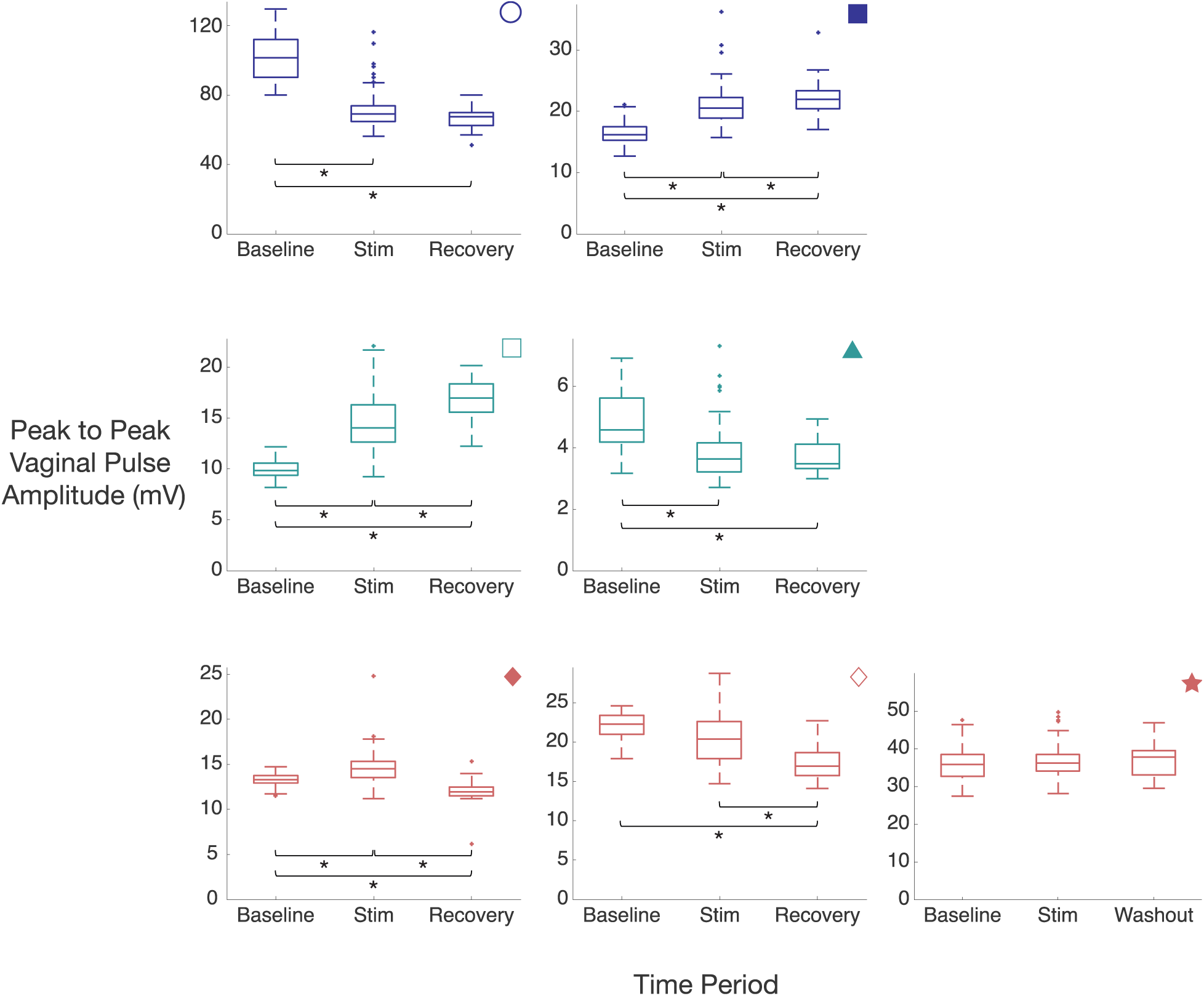
Boxplots of peak-to-peak VPA across different time periods (Baseline, Stim, and Recovery) in DGNS sessions. Boxplot *central lines* give the median, *edges* indicate the interquartile range, and *dots* represent outliers. Color and icons correspond to participants and groups as per Figure 2 and Table 2. * denotes significant difference (p < 0.05).

**Figure 4.**
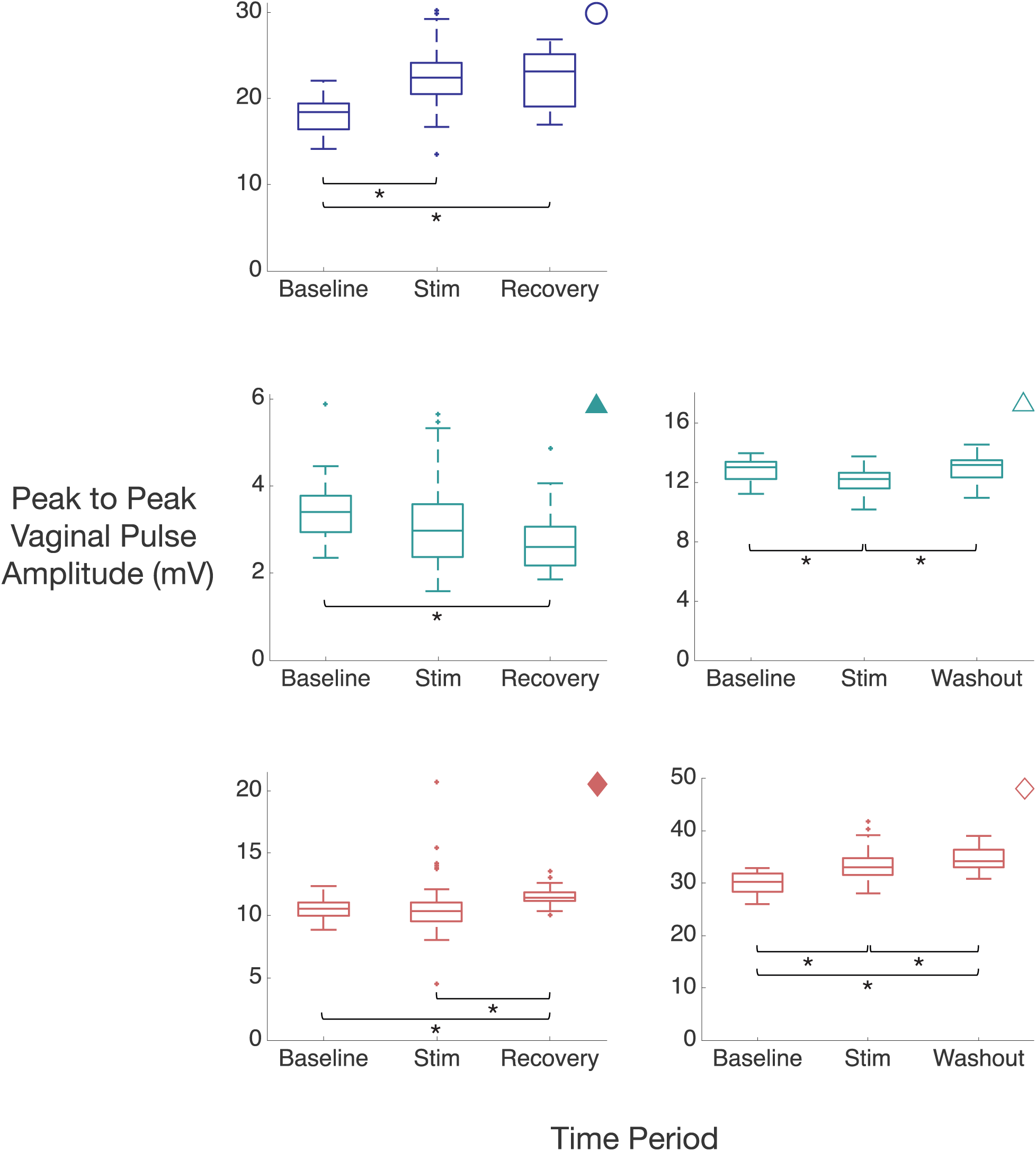
Boxplots of peak-to-peak VPA across different time periods (Baseline, Stim, and Recovery) in TNS sessions. Color and icons correspond to participants and groups as per Figure 2 and Table 2. * denotes significant difference (p < 0.05).

**Table 4.** Summary of VPA mean values (mV) during transcutaneous DGNS trials.

| Subject | Stimulation amplitude (mV) | Baseline (mean ± se) | Stimulation (mean ± se) | Recovery (mean ± se) | Baseline to Stimulation (%) | Baseline to Recovery (%) | Stimulation to Recovery (%) |
| --- | --- | --- | --- | --- | --- | --- | --- |
| NDAB-1 | 2 | No VPA data |  |  |  |  |  |
| NDAB-2 | 5.5 | 102.2 ± 2.7 | 70.6 ± 0.9 | 66.9 ± 1.1 | -30.9 | -34.5 | -5.3 |
| NDAB-3 | 3.5 | 16.4 ± 0.4 | 20.8 ± 0.3 | 22.2 ± 0.5 | +26.5 | +35.5 | +7.1 |
| SCI-1 | 60 | 10.0 ± 0.2 | 14.5 ± 0.3 | 17.0 ± 0.4 | +45.5 | +70.7 | +17.4 |
| SCI-2 | 60 | 4.9 ± 0.2 | 3.8 ± 0.1 | 3.7 ± 0.1 | -22.2 | -24.0 | -2.4 |
| FSD-1 | 16 | 13.2 ± 0.2 | 14.6 ± 0.2 | 12.0 ± 0.3 | +10.4 | -9.4 | -17.9 |
| FSD-2 | 60 | 22.0 ± 0.3 | 20.6 ± 0.3 | 17.5 ± 0.4 | -6.2 | -20.4 | -15.1 |
| FSD-3 | 23.5 | 36.1 ± 0.9 | 36.7 ± 0.4 | 37.6 ± 0.9 | +1.7 | +4.1 | +2.4 |
| mean ± se | 28.8 ± 9.5 | 29.2 ± 33.7 | 25.9 ± 22.1 | 25.3 ± 21.2 | +3.5 ± 26.7 | +3.1 ± 37.6 | -2.0 ± 12.3 |

## Discussion

In this preliminary investigation, we sought to understand the effect of one-time transcutaneous DGNS and TNS on genital arousal, measured as vaginal pulse amplitude. We also investigated the effect of DGNS and TNS on subjective arousal. This is the first study of its kind to measure genital arousal in response to neuromodulation in women. We observed a significant increase in subjective arousal during DGNS but not TNS across patients, and varying effects on VPA across stimulation sessions and patient groups. Women in all three participant groups gave positive comments to receiving acute neuromodulation and women with complete SCI experienced genital sensations in response to DGNS and TNS.

Subjective arousal increased following stimulation across all groups of participants in DGNS sessions. As expected, the FSD group had lower subjective arousal than their NDAB and SCI counterparts after stimulation. Sexual function involves complex coordination of the parasympathetic, sympathetic, and central nervous system. There are several different proposed models of the sexual response cycle^32–34^. Most models recognize that deficits in physiological areas (e.g., arousal) can influence the psychological (e.g., desire), making it difficult to pin-point the reason for lower subjective arousal in the participants with FSD. It is possible that the awareness of their FSD status in an isolated, clinical setting made them feel uncomfortable and led to lower subjective arousal. Participants with FSD experienced the fewest genital sensations and one participant (FSD-2) indicated that they might feel more receptive to using DGNS in a non-clinical setting (Table 3). Unexpectedly, some participants in DGNS sessions (3/8) had increases in subjective arousal before stimulation was turned on. This could perhaps be due to the presence of the VPA probe or anticipation of increased arousal.

All participants had significant changes in their peak-to-peak VPA between at least two periods (baseline, stim, or recovery) (Figure 3, Figure 4). Three participants (NDAB-3, FSD-1, and FSD-2) had opposing trends in their peak-to-peak VPA between DGNS and TNS sessions. It may be that not all women are responders to DGNS or TNS, as is common for neuromodulation therapies^35–37^. There were no consistent VPA trends (decrease, increase, no change) within groups, which is not entirely unexpected given the small sample size. The variability in our stimulation amplitudes (Table 4, Table 5) may have contributed to participants receiving sub-optimal current levels, dampening VPA responses. Either DGNS or TNS for sexual dysfunction may be most effective in women with reduced sensitivity to stimulation (e.g. our SCI cohort). However, we hypothesized that DGNS would increase VPA based on prior animal studies receiving stimulation at the pudendal nerve, the main trunk of the dorsal genital nerve. Cai et al^17^ theorized that pudendal nerve stimulation activates spinal autonomic efferents via the pelvic nerve. Larger studies are needed to examine these trends further. For example, in women with SCI, it could be that one nerve target is more effective than the other depending on their injury level and severity. However, this study is underpowered to make any claims of clinical efficacy for women with SCI.

**Table 5.** Summary of VPA mean values (mV) during transcutaneous TNS trials.

| Subject | Stimulation amplitude (mV) | Baseline (mean ± se) | Stimulation (mean ± se) | Recovery (mean ± se) | Baseline to Stimulation (%) | Baseline to Recovery (%) | Stimulation to Recovery (%) |
| --- | --- | --- | --- | --- | --- | --- | --- |
| NDAB-1 | 2 | No VPA data |  |  |  |  |  |
| NDAB-2 | 27.5 | 18.1 ± 0.4 | 22.5 ± 0.3 | 22.4 ± 0.6 | +24.3 | +23.7 | -0.5 |
| SCI-2 | 60 | 3.5 ± 0.1 | 3.1 ± 0.1 | 2.7 ± 0.7 | -11.1 | -21.0 | -11.1 |
| SCI-3 | 60 | 12.8 ± 0.2 | 12.1 ± 0.1 | 13.0 ± 0.1 | -5.0 | +1.6 | +7.0 |
| FSD-1 | 11 | 10.5 ± 0.2 | 10.4 ± 0.2 | 11.6 ± 0.1 | -0.9 | +9.9 | +10.8 |
| FSD-2 | 40 | 30.2 ± 0.4 | 33.3 ± 0.2 | 34.6 ± 0.4 | +10.4 | +14.6 | +3.8 |
| mean ± se | 33.4 ± 10.0 | 15.0 ± 10.0 | 16.3 ± 11.8 | 16.8 ± 12.1 | +3.5 ± 14.0 | +5.7 ± 17.0 | +2.0 ± 8.4 |

As participants were not exposed to sexual stimuli during the study procedure, our VPA results are not directly comparable with prior VPA studies. However, we believe this study provides important baseline data for VPA in the absence of erotic stimuli. A future study may seek to compare the effect of different types of erotic stimuli (e.g., audio, visual) on VPA to better understand genital arousal metrics. It is possible that repeated neuromodulation sessions over time, as is common for percutaneous tibial nerve stimulation for bladder dysfunction^38^, may yield consistent VPA changes. A future study may find more clinically meaningful results by examining the VPA response before and after multiple sessions of stimulation. Longitudinal VPA data may provide insights into a prior study that found repeated stimulation can increase FSFI scores^14^. Although animal studies have reported genital blood flow increases in response to tibial or pudendal nerve stimulation^15–18,39^, those studies used anesthetized animals and directly stimulated the nerve, which limits direct comparisons to this study. An awake, longitudinal animal model study, such as the one by Zimmerman et. al in which they found increased sexual receptivity after 6 weeks of biweekly TNS^40^, would provide a more analogous study paradigm to clinical studies and may find increased genital arousal.

Subjective arousal increased for one participant (SCI-2) in TNS sessions. It is likely that the mechanisms of TNS to modulate sexual function involve an indirect pathway to the pelvic organs, similar to the spinal reflexes proposed for bladder function^41^. Notably, this participant has a complete SCI (AIS-A) and reports that she does not experience any sensation below her level of injury (T5). It is possible that there are residual fibers in her spinal cord that are carrying afferents that were activated by DGNS or TNS. Another hypothesis is that afferents from the genitals during arousal, activated by DGNS or TNS, could be circumventing the spinal cord via the vagus nerve, leading to subjective arousal. One study in women with complete SCI asked participants to perform vaginal-cervical self-stimulation in a functional magnetic resonance imaging machine. Researchers found that the nucleus solitary tract (NTS), where the vagus nerve projects in the brain stem, was active during self-stimulation^42^. This is supported by an animal study that found neurons in the NTS that responded to vaginal distension and cervical stimulation^43^.

Almost all women were willing to use DGNS or TNS outside of this study, provided it would help with their sexual dysfunction (if they had it). There was more hesitancy with DGNS as a therapy among the FSD group, perhaps due to the sensitive location of the electrodes. All participants who were lost to follow up after their first study session received DGNS, which may suggest that the method for delivering DGNS could be improved. Both nerve targets have distinct advantages. DGNS was able to increase subjective arousal but is applied at a sensitive location. Women may find they are more comfortable with stimulation at their ankle rather than their genitals and TNS is easier to administer at the ankle, however the mechanisms of how it improves sexual function are less clear.

Our study did not include any audio-visual materials, so we are limited in comparing our results to clinical arousal studies that include videos with their interventions^44^. Another limitation of our study is that we did not confirm electrode placement for TNS or DGNS. A future study may confirm target nerve recruitment by verifying activation of a motor response (e.g. toe twitch) or pudendo-anal reflex for TNS and DGNS respectively. An additional consideration may be to place one electrode directly over the clitoris and a return electrode on the labia majora or inner thigh as in Bourbeau et al.^45^. This configuration may provide more targeted stimulation of the genital nerve as it travels along the crura of the clitoris.

We are also limited by our sample size (n = 3 per group), which prevented us from identifying any conclusive trends in genital arousal within or across participant groups. Unfortunately, this sample size is too small to make any claims of clinical efficacy, but we have now established a baseline for VPA response to DGNS and TNS in a one-time session. Although there were no trends in VPA across participants, individual participants had a VPA_Change_ as large as +70.7%. We believe that these are important values to report, despite the small sample size, because it provides important context for a future study to add erotic stimuli to stimulation sessions. It is likely that the clinical environment with study team members present coupled with the lack of sexual stimuli contributed to an inhibition of sexual arousal, dampening genital arousal responses.

## Conclusion

To our knowledge, this is the first clinical study to measure subjective and genital arousal in response to acute neuromodulation. We sought to understand the impact of two potential treatment modalities, transcutaneous DGNS and TNS, on subjective arousal and vaginal pulse amplitude during a one-time neuromodulation session. We found that DGNS, but not TNS, increased subjective arousal across all participants. We did not observe a consistent VPA response to DGNS or TNS across all participants or within participant groups. All SCI participants experienced genital sensations during DGNS and TNS sessions, although given the small sample size, a larger study is warranted before making claims of clinical efficacy. Future studies may incorporate audio-visual materials or another type of sexual stimuli to better facilitate arousal. Studies with repeated stimulation sessions over time may find more clinically relevant improvements in sexual function.

## Data Availability

All data produced are available online at OSF.IO/G7FQ9.

## Acknowledgements

We thank Vanessa Pruitt for initial assistance with study recruitment, the Michigan Clinical Research Unit, Patricia Maymi-Castrodad, and Zhina Sadeghi for study session support, and Chris Andrews for help with statistical analysis. This study was supported by the Craig H. Neilsen Foundation (grant number 647332), the National Institutes of Health Award T32NS115724, and the International Society for the Study of Women’s Sexual Health.

## References

1. Nappi RE, Cucinella L, Martella S, Rossi M, Tiranini L, Martini E. Female sexual dysfunction (FSD): Prevalence and impact on quality of life (QoL). Maturitas. 2016;94:87–91. doi:10.1016/j.maturitas.2016.09.013

2. Dhillon S, Keam SJ. Bremelanotide: First Approval. Drugs. 2019;79(14):1599–1606. doi:10.1007/s40265-019-01187-w

3. Dooley EM, Miller MK, Clayton AH. Flibanserin: From Bench to Bedside. Sex Med Rev. 2017;5(4):461–469. doi:10.1016/j.sxmr.2017.06.003

4. Basson R, McInnes R, Smith MD, Hodgson G, Koppiker N. Efficacy and Safety of Sildenafil Citrate in Women with Sexual Dysfunction Associated with Female Sexual Arousal Disorder. J Womens Health Gend Based Med. 2002;11(4).

5. Anderson KD. Targeting recovery: Priorities of the spinal cord-injured population. J Neurotrauma. 2004;21(10):1371–1383. doi:10.1089/neu.2004.21.1371

6. Collinger JL, Boninger ML, Bruns TM, Curley K, Wang W, Weber DJ. Functional priorities, assistive technology, and brain-computer interfaces after spinal cord injury. J Rehabil Res Dev. 2013;50(2):145–160. doi:10.1682/JRRD.2011.11.0213

7. Kashdan TB, Goodman FR, Stiksma M, Milius CR, McKnight PE. Sexuality leads to boosts in mood and meaning in life with no evidence for the reverse direction: A daily diary investigation. Emotion. 2018;18(4):563–576. doi:10.1037/emo0000324

8. Krassioukov A, Elliott S. Neural control and physiology of sexual function: Effect of spinal cord injury. Top Spinal Cord Inj Rehabil. 2017;23(1):1–10. doi:10.1310/sci2301-1

9. Rosen R, Brown C, Heiman J, et al. The female sexual function index (Fsfi): A multidimensional self-report instrument for the assessment of female sexual function. J Sex Marital Ther. Published online 2000. doi:10.1080/009262300278597

10. Lombardi G, Mondaini N, Macchiarella A, Cilotti A, Popolo GD. Clinical Female Sexual Outcome after Sacral Neuromodulation Implant for Lower Urinary Tract Symptom (LUTS). J Sex Med. 2008;5(6):1411–1417. doi:10.1111/j.1743-6109.2008.00812.x

11. Yih JM, Killinger KA, Boura JA, Peters KM. Changes in Sexual Functioning in Women after Neuromodulation for Voiding Dysfunction. J Sex Med. 2013;10(10):2477–2483. doi:10.1111/jsm.12085

12. Banakhar MA, Gazwani Y, ElKelini M, Al-shaiji T, Hassouna M. Effect of sacral neuromodulation on female sexual function and quality of life: Are they correlated? Can Urol Assoc J. 2014;8(11-12):762. doi:10.5489/cuaj.2300

13. Kershaw V, Khunda A, McCormick C, Ballard P. The effect of percutaneous tibial nerve stimulation (PTNS) on sexual function: a systematic review and meta-analysis. Int Urogynecology J. 2019;30(10):1619–1627. doi:10.1007/s00192-019-04027-3

14. Zimmerman LL, Gupta P, O’Gara F, Langhals NB, Berger MB, Bruns TM. Transcutaneous Electrical Nerve Stimulation to Improve Female Sexual Dysfunction Symptoms: A Pilot Study. Neuromodulation. 2018;21(7):707–713. doi:10.1111/ner.12846

15. Zimmerman LL, Rice IC, Berger MB, Bruns TM. Tibial Nerve Stimulation to Drive Genital Sexual Arousal in an Anesthetized Female Rat. J Sex Med. 2018;15(3):296–303. doi:10.1016/j.jsxm.2018.01.007

16. Xu JJ, Zimmerman LL, Soriano VH, et al. Tibial nerve stimulation increases vaginal blood perfusion and bone mineral density and yield load in ovariectomized rat menopause model. Int Urogynecology J. Published online 2022. doi:10.1007/s00192-022-05125-5

17. Cai RS, Alexander MS, Marson L. Activation of Somatosensory Afferents Elicit Changes in Vaginal Blood Flow and the Urethrogenital Reflex Via Autonomic Efferents. J Urol. 2008;180(3):1167–1172. doi:10.1016/j.juro.2008.04.139

18. Rice IC, Zimmerman LL, Ross SE, Berger MB, Bruns TM. Time-Frequency Analysis of Increases in Vaginal Blood Perfusion Elicited by Long-Duration Pudendal Neuromodulation in Anesthetized Rats. Neuromodulation. 2017;20(8):807–815. doi:10.1111/ner.12707

19. Maroufizadeh S, Riazi H, Lotfollahi H, Omani-Samani R, Amini P. The 6-item Female Sexual Function Index (FSFI-6): factor structure, reliability, and demographic correlates among infertile women in Iran. Middle East Fertil Soc J. 2020;24(1):7. doi:10.1186/s43043-019-0008-8

20. Scarpero HM, Fiske J, Xue X, Nitti VW. American Urological Association Symptom Index for lower urinary tract symptoms in women: Correlation with degree of bother and impact on quality of life. Urology. 2003;61(6):1118–1122. doi:10.1016/S0090-4295(03)00037-2

21. Rockwood TH, Church JM, Fleshman JW, et al. Patient and surgeon ranking of the severity of symptoms associated with fecal incontinence: The fecal incontinence severity index. Dis Colon Rectum. 1999;42(12):1525–1531. doi:10.1007/BF02236199

22. Frank L, Kleinman L, Farup C, Taylor L, Miner P. Psychometric validation of a constipation symptom assessment questionnaire. Scand J Gastroenterol. 1999;34(9):870–877. doi:10.1080/003655299750025327

23. Ware J, Sherbourne C. The MOS 36-Item Short-Form Health Survey (SF-36): I. Conceptual Framework and Item Selection. Med Care. 2012;30(6):473–483.

24. Harris PA, Taylor R, Thielke R, Payne J, Gonzalez N, Conde JG. Research electronic data capture (REDCap)-A metadata-driven methodology and workflow process for providing translational research informatics support. J Biomed Inform. 2009;42(2):377–381. doi:10.1016/j.jbi.2008.08.010

25. Prause N, Janssen E. Blood flow: Vaginal photoplethysmography. In: Women’s Sexual Function and Dysfunction: Study, Diagnosis and Treatment.; 2006:361–369.

26. Opisso E, Borau A, Rijkhoff NJM. Subject-Controlled Stimulation of Dorsal Genital Nerve to Treat Neurogenic Detrusor Overactivity at Home. Neurourol Urodyn. 2013;32:1004–1009. doi:10.1002/nau

27. Díaz-Ruiz MDC, Romero-Galisteo RP, Arranz-Martín B, Palomo-Carrión R, Ando-Lafuente S, Lirio-Romero C. Vibration or Transcutaneous Tibial Nerve Stimulation as a Treatment for Sexual Dysfunction in Women with Spinal Cord Injury: Study Protocol for a Randomized Clinical Trial. Int J Environ Res Public Health. 2022;19(3):1478. doi:10.3390/ijerph19031478

28. Prause N, Williams K, Bosworth K. Wavelet denoising of vaginal pulse amplitude. Psychophysiology. 2010;47(2):393–401. doi:10.1111/j.1469-8986.2009.00941.x

29. Pulverman CS, Meston CM, Hixon JG. Automated Artifact-Detection Procedure for Vaginal Photoplethysmography. J Sex Marital Ther. 2018;44(6):566–590. doi:10.1080/0092623X.2018.1436627

30. Prause N, Barela J, Roberts V, Graham C. Instructions to Rate Genital Vasocongestion Increases Genital and Self-Reported Sexual Arousal but not Coherence Between Genital and Self-Reported Sexual Arousal. J Sex Med. 2013;10(9):2219–2231. doi:10.1111/jsm.12228

31. Bottorff E, Bruns T. Acute dorsal genital nerve stimulation increases subjective arousal in women with and without spinal cord injury. Published online 2023. doi:10.17605/OSF.IO/G7FQ9

32. Pfaus JG. Pathways of Sexual Desire. J Sex Med. 2009;6(6):1506–1533. doi:10.1111/j.1743-6109.2009.01309.x

33. Kingsberg SA, Janata JW. Female Sexual Disorders: Assessment, Diagnosis, and Treatment. Urol Clin North Am. 2007;34(4):497–506. doi:10.1016/j.ucl.2007.08.016

34. Basson R. The Female Sexual Response: A Different Model. J Sex Marital Ther. 2000;26(1):51–65. doi:10.1080/009262300278641

35. Blok B, Van Kerrebroeck P, de Wachter S, et al. Three month clinical results with a rechargeable sacral neuromodulation system for the treatment of overactive bladder. Neurourol Urodyn. 2018;37(S2):S9–S16. doi:10.1002/nau.23465

36. Khan H, Pilitsis JG, Prusik J, Smith H, McCallum SE. Pain Remission at One-Year Follow-Up With Spinal Cord Stimulation. Neuromodulation Technol Neural Interface. 2018;21(1):101–105. doi:10.1111/ner.12711

37. Hödl S, Carrette S, Meurs A, et al. Neurophysiological investigations of drug resistant epilepsy patients treated with vagus nerve stimulation to differentiate responders from non-responders. Eur J Neurol. 2020;27(7):1178–1189. doi:10.1111/ene.14270

38. Peters KM, Carrico DJ, Perez-Marrero RA, et al. Randomized Trial of Percutaneous Tibial Nerve Stimulation Versus Sham Efficacy in the Treatment of Overactive Bladder Syndrome: Results From the SUmiT Trial. J Urol. 2010;183(4):1438–1443. doi:10.1016/j.juro.2009.12.036

39. Bottorff EC, Bruns TM. Pudendal, but not tibial, nerve stimulation modulates vulvar blood perfusion in anesthetized rodents. Int Urogynecology J. Published online November 3, 2022. doi:10.1007/s00192-022-05389-x

40. Zimmerman LL, Mentzelopoulos G, Parrish H, et al. Immediate and Long-Term Effects of Tibial Nerve Stimulation on the Sexual Behavior of Female Rats. Neuromodulation Technol Neural Interface. Published online January 2023:S109471592201371X. doi:10.1016/j.neurom.2022.11.008

41. de Groat WC, Tai C. Mechanisms of Action of Sacral Nerve and Peripheral Nerve Stimulation for Disorders of the Bladder and Bowel. In: Neuromodulation. Elsevier; 2018:221–236. doi:10.1016/B978-0-12-805353-9.00019-X

42. Komisaruk BR, Whipple B. Functional MRI of the Brain During Orgasm in Women. Annu Rev Sex Res. 2005;16(1):62–86. doi:10.1080/10532528.2005.10559829

43. Hubscher CH, Berkley KJ. Responses of neurons in caudal solitary nucleus of female rats to stimulation of vagina, cervix, uterine horn and colon. Brain Res. 1994;664(1-2):1–8. doi:10.1016/0006-8993(94)91946-1

44. Handy AB, Stanton AM, Pulverman CS, Meston CM. Differences in Perceived and Physiologic Genital Arousal Between Women With and Without Sexual Dysfunction. J Sex Med. 2018;15(1):52–63. doi:10.1016/j.jsxm.2017.11.009

45. Bourbeau DJ, Creasey GH, Sidik S, Brose SW, Gustafson KJ. Genital nerve stimulation increases bladder capacity after SCI: A meta-analysis. J Spinal Cord Med. 2018;41(4):426–434. doi:10.1080/10790268.2017.1281372

46. Wiegel M, Meston C, Rosen R. The Female Sexual Function Index (FSFI): Cross-Validation and Development of Clinical Cutoff Scores. J Sex Marital Ther. 2005;31(1):1–20. doi:10.1080/00926230590475206

